# Application of self-reporting, clinicians’ assessment, and tacrolimus trough levels in evaluating adherence to immunosuppressive therapy in kidney transplant recipients. A cohort study

**DOI:** 10.1101/2023.08.14.23294064

**Authors:** Choki Dorji, Tashi Tobgay, Kesara Na-Bangchang

**Affiliations:** Graduate Program in Bioclinical Sciences, Chulabhorn International College of Medicine, Thammasat University (Rangsit Campus), Pathum Thani, Thailand; Department of Pharmacy, Jigme Dorji Wangchuk National Referral Hospital, Thimphu Bhutan; Institute of Health Partners, Phendey lam, Thimphu, Bhutan; Drug Discovery and Development Center, Office of Advanced Science and Technology, Thammasat University (Rangsit Campus), Pathum Thani, Thailand

**Keywords:** kidney transplant, adherence, nonadherence, tacrolimus, kidney function, immunosuppressive therapy, Therapeutic drug monitoring

## Abstract

**Introduction:** Immunosuppressants are drugs with narrow therapeutic indices and pharmacokinetic variation. Nonadherence to the therapy will cause over or underexposure leading to graft rejection.

**Methods:** A cohort study was conducted on kidney transplant recipients from the nephrology clinic and therapeutic drug monitoring unit. Patients were asked to self-report their medication adherence using a self-assessment tool. Assessment by clinicians and variation of tacrolimus levels were evaluated. Tacrolimus concentration and kidney function were measured prospectively to correlate with patients’ adherence. The variation of tacrolimus levels between 20-43 % was classified as medium and above 43 % as poor adherence.

**Results:** Among 58 participants, 33 (56.7%) were females. The maximum number of years attained after transplantation was 17, with a median duration of 5 years. On the self-reporting scale, 47.0% of adherence was due to fear of graft rejection. Among nonadherences, 77.4% had their immunosuppressive therapy two hours before or after the prescribed time. Based on the clinician score, 39 (67.2%) and 42 (72.4%) cases were identified as nonadherence and having tacrolimus C _trough_ level above 20 % respectively. The median (95% CI) serum creatine and blood urea nitrogen levels in the medium category were 1.23 mg/dL (1.2-1.4) (P = 0.009) and 28.3 mg/dL (26.4-36.4) (P = 0.021) respectively. The corresponding values for poor adherence were 2.5 mg/dL (1.6-3.5) (P = 0.03) and 43.0 mg/dL (35.5-78.0) (P = 0.01).

**Conclusions:** The fear of allograft rejection is linked to a better adherence rate. Nonadherent transplant recipients required close observation and frequent monitoring of immunosuppressant levels for graft survival.

## Introduction

Patients undergoing solid organ transplants are obligated to adhere to a lifelong immunosuppressive therapy (IST) tailored to preserve long-term graft function. Advancing surgical techniques and the introduction of novel IST have prolonged the survival rate of up to five years of organ transplants to 88.0 %. (1) Unsurprisingly, nonadherence to IST diverging from posttransplant medical advice has repeatedly been demonstrated to predict morbidity and mortality in organ transplant recipients. (2) In kidney transplant recipients (KTRs), nonadherence to IST is as high as 35.6 % annually. This incidence is higher than that of the general population of solid organ transplant recipients (22.6 % annually). (3) The WHO defines adherence as the extent to which the person’s action, such as taking medication, following a diet and /or executing lifestyles, corresponds with agreed recommendations from a healthcare provider. (4) It is estimated that approximately 30-50% of the prescribed IST are not taken as directed (5), indicating nonadherence as the cause of graft loss, infection, healthcare expense, and poor quality of life (QoL). (6–8) Nonadherence occurs when the patient administers the drug without complying with what is documented in the clinical chart, strong clinical suspicion by the attending clinicians, subtherapeutic immunosuppressive levels, and repeated failure of clinic visits. (9) Considering these factors, the consensus report defines nonadherence as a deviation from the prescribed medication regimen sufficient to influence the intended effect adversely. (10) The WHO broadly correlates factors associated with nonadherence as socioeconomic, healthcare systems, treatment condition, and patient-related factors. (11) Evidence shows a gradual decline in adherence with prolonged length of posttransplant duration, resulting in a higher incidence of late acute rejection among nonadherence. (12,13) Like in any other condition, medication nonadherence in KTRs increases with the number of medications prescribed. (14) Social support plays a role in the adherence rate, with married people showing higher levels of adherence than single individuals. (15) According to some research, employees exhibit better adherence rates than unskilled employees, while other studies find no difference in adherence regardless of socioeconomic class or educational attainment. (13) Low income did not impact medication adherence but was linked to worse kidney allograft survival. (16) Despite being aware of the significance of IST, the majority do not adhere for a variety of reasons, including medication side effects, dosage frequency, and complications. (17) Forgetfulness was reported as a contributing factor to poor adherence among people between the ages of 18 and 29 years in the United States. (5) According to the report, adolescents reported lower adherence rates than adults. (6) The rates and factors associated between gender and nonadherence are inconsistent. (18) More than 25% of patients enrolled in IST clinical trials were found as nonadherence. (7)

There are several direct and indirect methods for assessing medication adherence. Each has benefits and drawbacks that must be balanced against the accuracy, expense, and burden involved. (8) Monitoring serum drug levels is an essential predictor for drug toxicity, dose adjustment, and adherence. (19) Although self-reports should continue to be the mainstay of adherence assessment, there should be attention given to other approaches to identify medication nonadherence. (20) Most studies on medication adherence in transplant patients are focused on immunosuppressive medication because of its impact on preventing acute graft rejection. The methods available for testing tacrolimus concentration provide an objective measurement of nonadherence. Other studies were conducted using self-reporting tools or pill counts. Nevertheless, patients generally underestimate medication nonadherence through this method, so the incidence of nonadherence in renal transplant recipients is likely higher than generally appreciated. (21) In this study, we intend to address the level of adherence to tacrolimus IST by applying three indicators, *i.e*., self-reporting by applying the Basel Assessment of Adherence to Immunosuppressive Medication Scale (BAASIS) tool, clinicians’ assessment, and variation of tacrolimus trough concentration.

## Materials & Methods

### Settings

The study was conducted during January and October 2022, at the Jigme Dorji Wangchuk national referral hospital, a tertiary care hospital. This is the only hospital that provides nephrology and therapeutic drug monitoring (TDM) services in Bhutan. An observational study was conducted among KTRs undergoing surgery until December 2021.

### Study design and population

We recruited KTRs who came for nephrology consultation and tacrolimus level monitoring. The nature of the study was explained to all participants, and their participation was voluntary. Patients with a chronic condition and active infection requiring admission and monitoring of the tacrolimus area under the concentration curve (AUC) were excluded from the study.

### Data collection and measurement tool

The data collection tool comprises sociodemography information, a questionnaire on adherence (self-reporting), clinicians’ assessment, and laboratory data - Tacrolimus C _trough_, serum creatinine, and blood urea nitrogen (BUN). The information on the quality of life (QoL) of the participants was obtained from the previous study. (22)

#### Adherence measurement

Primary assessment using a self-reporting BAASIS scale was used to capture the medication taking and timing profile. The questionnaire was administered in English and Dzongkhag (Bhutan National language) versions after validation by the experts. A verbal explanation was given to those having difficulty in reading. The BAASIS tool is a proper self-reporting instrument that consists of four items which assess adherence to IST over four weeks. This instrument assesses four dimensions of adherence, i.e., missing dose, continuous missing of several doses, deviation from the exact medication taking time for longer than two hours, and reducing the amount of each dose. The responses were ranked in the order of a 6-point Likert rating scale from 0 to 5 (0 = never, 1 = once a month, 2 = every 2 weeks, 3 = every week, 4 = more than once a week, and 5 = every day). (23) The assessment tool was piloted for content validity by the experts and translated back into Dzongkhag. Discrepancies were removed and made easily understandable in the local context. Cronbach’s alpha was used to determine the internal consistency. A value greater than or equal to 0.78 was considered acceptable.

Secondary assessment was based on the clinician’s view on adherence, i.e., the frequency of past missing appointments, clinical tests, and inappropriate dosing. The adherence was categorized as “good” if any of the parameters missed was less than one time, “medium” if the parameters missed were between one and three times, and “poor” if the parameters missed were more than four times.

The tertiary assessment was based on tacrolimus C_trough_ levels. Tacrolimus whole□blood samples were used to determine the 12-hour tacrolimus C_trough_ levels collected as part of the patient’s clinical care. Blood was collected before the morning dose. Tacrolimus concentrations were measured using the electrochemiluminescence immunoassay (ECLIA). The coefficient of variation (%CV) of tacrolimus C_trough_ was calculated using at least five tacrolimus C_trough_ levels using the formula %CV = (σ/μ) x 100 (σ=SD and μ=mean tacrolimus C_trough_. (24) Other variables included serum creatinine and BUN. The level of adherence was defined as “good” for CV < 20%, “medium” for CV 20-43%, and “poor” for CV > 43%.(25) The association between increased serum creatinine and BUN levels in nonadherence KTRs was assessed.

### Statistical analysis

Data collected were checked for normal distribution using Kolmogorov-Smirnov test, histogram, and Q-Q plots. Qualitative data are summarized as number (%) and quantitative data are summarized as mean ± SD, or median (95%CI), where appropriate. Comparisons between the %CV of tacrolimus, Trough levels, and serum creatinine and BUN were performed using Kruskal-Wallis test. Cohen’s kappa was applied to determine the level of agreement with the clinician’s score, and crosstabs were used to investigate the overlap. For group comparison between the BAASIS adherence scale and demographic information with continuous and categorical data, independent sample t-test, Mann-Whitney U test, and Pearson’s chi-square test were used, respectively. The statistical significance level was set at α = 0.05.

## Results

The median (range) age of 58 participants (56.7 % females) was 41 (23 – 65) years. Seventy-seven percent were married. The maximum number of years attained after the posttransplant was 17, with a median duration of 5 years. The information on employment status, education level, and IST is shown in Table 1. On a self-reporting scale, 27 (47.0%) recipients were identified as adherence, which was significantly linked to fear/uncertainty of allograft rejection [mean ± (SD): 4.5 ± (0.7), (P= 0.04)] on a Likert scale. The sociodemographic factors affecting adherence are shown in Table 2. Among nonadherence recipients, 24 (77.4%) took their medication two hours from the prescribed time. The frequency of missed dose deviation and reduced dose are shown in Table 3. Overall, there are forty-two transplant recipients having coefficient variation (%CV) greater than 20 % [mean ± SD: 29.96 ± 16.5 %] of tacrolimus C_trough_ levels, associated with increased serum creatine and BUN (Figure 1). The median (95% CI) values of serum creatine and BUN levels for % CV between 20 - 43 % were 1.23 (1.2-1.4) mg/dL (P=0.009) and 28.3 (26.4-36.4) mg/dL (P=0.021), respectively. The corresponding values for the % CV ≥ 43 % were 2.5 (1.6-3.5) mg/dL (P=0.03) and 43.0 (35.5-78.0) mg/dL (P=0.01), respectively. The kappa assessment of the clinician’s score did not differ from the tacrolimus C_trough_ levels, Table 4.

**Table 1:** Frequency distribution of sociodemographic distribution.

| **Age Range** | **Frequency (%) N=58** |
| --- | --- |
| 20-30 | 6(10.3%) |
| 30-40 | 22(37.9%) |
| 40-50 | 18(31.0%) |
| 50-60 | 7(12.1%) |
| 60-70 | 5(8.6%) |
| Max | 65 |
| Min | 23 |
| Median | 41 |
| **Sex** |  |
| Male | 25(43.1%) |
| Female | 33(56.9%) |
| **Marital status** |  |
| Married | 45(77.6%) |
| Never Married | 11(19.0%) |
| Divorce | 2(3.4%) |
| **Employment** |  |
| Employed | 28(48.3%) |
| Unemployed | 30(51.7%) |
| **Education level** |  |
| No formal education | 14(24.1%) |
| School | 25(43.1%) |
| University graduates | 14(24.1%) |
| Monastic education | 5(8.6%) |
| **Duration after transplant** |  |
| 0-5 years | 32(55.2%) |
| 5-10 years | 23(39.7%) |
| 10-20 years | 3(5.2%) |
| Max | 17 |
| Min | 0 |
| Median | 5 |
| **Immunosuppressant drugs** |  |
| Tacrolimus + Mycophenolate + Prednisolone | 49(84.5%) |
| Tacrolimus + Everolimus + Prednisolone | 6(10.3%) |
| Tacrolimus + Azathioprine | 2(3.4%) |
| Tacrolimus | 1(1.7%) |

**Table 2:** Sociodemographic factors affecting self-reporting BAASIS tool.

|  | **Adherence**<br>**N=27** | **Non-adherence**<br>**N=31** | **P-value** |
| --- | --- | --- | --- |
| Age years (mean ± SD) | 45 ± (11) | 41 ± (10) | 0.47<sup>†</sup> |
| **Sex** |  |  |  |
| Male | 10(37.0%) | 15(48.4%) | 0.43<sup>‡</sup> |
| Female | 17(63.0%) | 16(51.6%) |  |
| **Marital status** |  |  |  |
| Married | 23(85.2%) | 22(71.0%) | 0.31<sup>‡</sup> |
| Never Married | 3(11.1%) | 8(25.8%) |  |
| Divorce | 1(3.7%) | 1(3.2%) |  |
| **Occupation** |  |  |  |
| Employed | 11(40.7%) | 17(54.8%) | 0.20<sup>‡</sup> |
| Unemployed | 16(59.3%) | 14(45.2%) |  |
| **Education** |  |  |  |
| No formal education | 6(22.2%) | 8(25.8%) | 0.43<sup>‡</sup> |
| School | 14(51.9%) | 11(35.5%) |  |
| University graduates | 6(22.2%) | 8(25.8%) |  |
| Monastic education | 1(3.7%) | 4(12.9%) |  |
| Years after transplant (mean ± SD) | 5.11 ± (4.1) | 4.7 ± (2.7) | 0.11<sup>†</sup> |
| **Quality of Life dimensions<sup>¶</sup>** |  |  |  |
| Physical symptom (median) | 4.6 | 5.0 | 0.72<sup>§</sup> |
| Fatigue (mean ± SD) | 3.9 ± (1.1) | 4.5 ± (0.9) | 0.67<sup>†</sup> |
| Uncertainty/Fear (mean ± SD) | 4.3 ± (1.0) | 4.5 ± (0.7) | <0.05<sup>†</sup> |
| Appearance (mean ± SD) | 3.8 ± (1.4) | 4.0 ± (1.1) | 0.20<sup>†</sup> |
| Emotional (mean ± SD) | 3.9 ± (1.4) | 4.3 ± (1.4) | 0.75<sup>†</sup> |

Independent sample *t*-test<sup>†</sup>, Chai-square test<sup>‡</sup>, Mann-Whitney test<sup>§</sup>.
<sup>¶</sup>Quality of life (QoL) data was extracted from the previously conducted study of participants; lower the number better the QoL.

**Table 3:** Frequency of missing dose and time deviation from the exact medication-taking timing.

| **Times** | **Response** | **N (%)** | **Total** |
| --- | --- | --- | --- |

|  |  |  | (n=31) |
| --- | --- | --- | --- |
| Missed IST in the last 4 weeks | Once per month | 15(48.3%) | 19(61.3) |
|  | Once per 2 weeks | 1(3.2%) |  |
|  | Every week | 1(3.2%) |  |
|  | More than once per week | 2(6.4%) |  |
|  | Every day | 0(0.0%) |  |
| Several consecutive doses of IST in the last 4 weeks. | Once per month | 6(19.3%) | 14(45.1) |
|  | Once per 2 weeks | 4(12.9%) |  |
|  | Every week | 0(0.0%) |  |
|  | More than once per week | 3(9.6%) |  |
|  | Every day | 2(6.4%) |  |
| IST 2 hours before or after my prescribed time in the last 4 weeks | Once per month | 16(51.6%) | 24(77.4) |
|  | Once per 2 weeks | 2(6.4%) |  |
|  | Every week | 1(3.2%) |  |
|  | More than once per week | 5(16.1%) |  |
|  | Every day | 0(0.0%) |  |
| Lower dose of IST than the prescribed dose in the last 4 weeks | Once per month | 6(19.3%) | 8(25.8) |
|  | Once per 2 weeks | 1(3.2%) |  |
|  | Every week | 0(0.0%) |  |
|  | More than once per week | 1(3.2%) |  |
|  | Every day | 0(0.0%) |  |

**Table 4:** Overlapping score between clinicians’ assessment and tacrolimus (C _trough_) concentration. Data are presented as number (%) values.

| | | Variation of tacrolimus ($C_{\text{trough}}$) concentration | | | P-Value |
| --- | --- | --- | --- | --- | --- |
|  |  | Good (n=16) | Medium (n=33) | Poor (n=9) |  |
| **Clinicians' score** | Good (n=19) | 11(68.7%) | 8(24.2%) | 0(0.0%) | <0.001* |
|  | Medium (n=34) | 5(31.2%) | 24(72.7%) | 5(55.5%) |  |
|  | Poor (n=5) | 0(0.0%) | 1(3.0%) | 4(44.4%) |  |

\* The Kappa assessment score by the clinician does not differ significantly from CV % at 0.96 ( $\alpha=0.05$ ) and the level of agreement is Excellence (0.93-1.00).
Adherence based on percentage covariate of variation (CV%) is classified as good if CV % = < 20 %, medium for CV = 20-43%, and CV = > 43% as poor.

**Figure 1:**
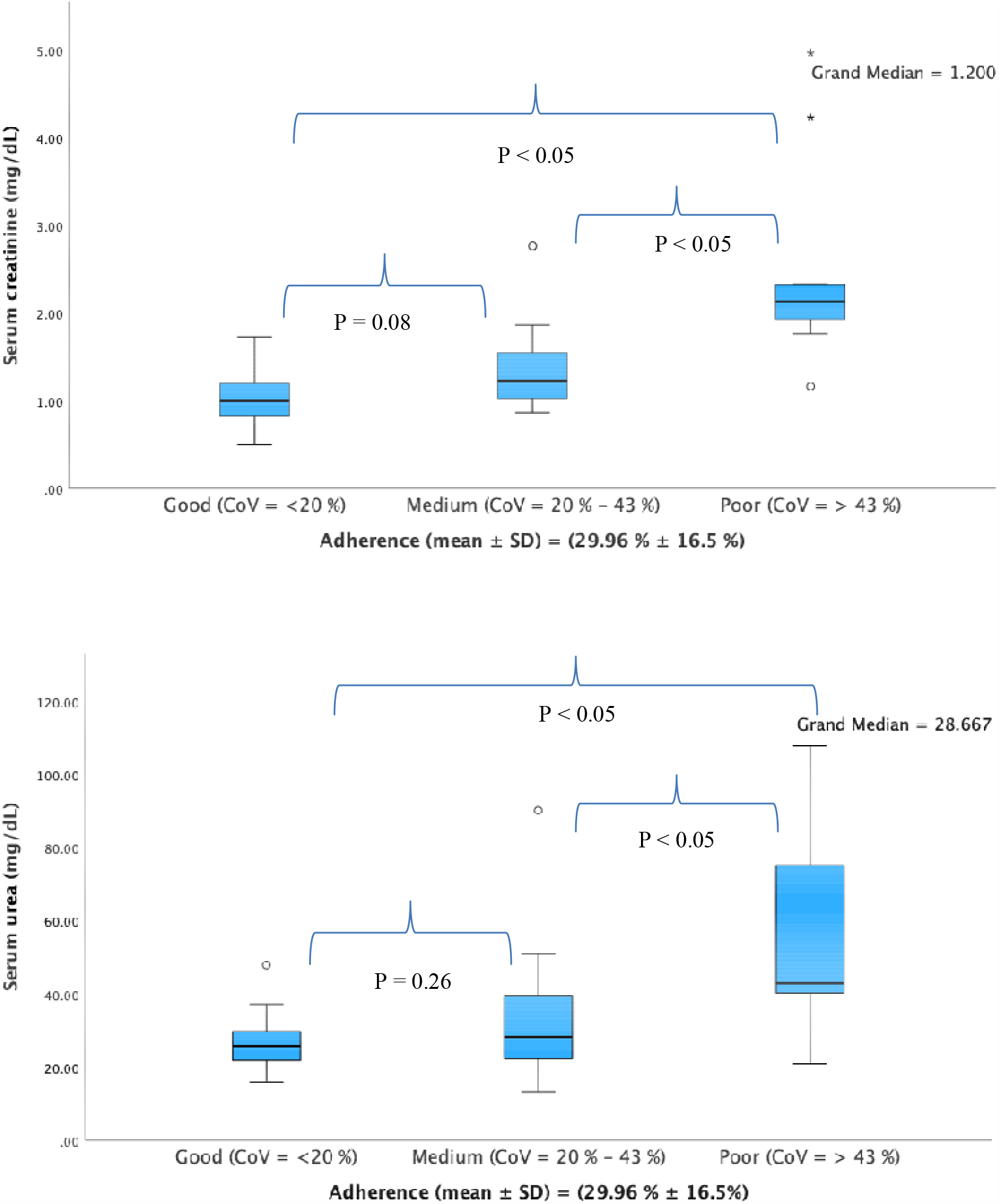
% CV of tacrolimus C _trough_ vs serum creatinine and BUN.

## Discussion

There is robust evidence to support the use of various techniques and tools in assessing adherence to IST. This study was conducted to assess the rate of adherence in Bhutanese KTRs by applying three different methods. This study is significant to Bhutan because there is a lack of kidney transplant centers in Bhutan. Patients requiring surgery are referred to India by the state, and the state covers the cost of referral and life-long maintenance of IST after transplantation. As enshrined in Section 21 of Article 9 of the constitution, it is mentioned that “The State shall provide free access to basic public health services in both modern and traditional medicine”.(26) Kidney transplantation is a life-saving procedure for individuals with end-stage renal disease, offering better QoL and increased survival rates. The IST plays a vital role in ensuring the success of kidney transplantation by preserving the kidney from allograft rejection. However, nonadherence to IST poses a significant risk of rejection and adverse outcomes.

Our study found 53.5% of the study participants were not adhering to IST on the self-reporting scale. The nonadherence was identified based on 39 (67.2%) clinicians’ scores, and 42 (72.4%) with %CV of tacrolimus C_trough_ level >20%. The self-reporting scale is based on the reporter’s perception used for assessing the factors related to the QoL after transplantation (27) and results showed a significant link between adherence and fear/uncertainty of the graft rejection domain of QoL. Similar studies also showed psychological disorders as predictors of nonadherence. (20) Such problems can be efficiently treated to improve QoL, which is crucial if adherence is to be improved. The present study’s adherence rate was consistent with previous studies results. (28) Among nonadherence, 24 (77.4%) participants reported taking IST 2 hours before or after the prescribed time, and 19 (61.3 %) participants missed at least one dose in a month. To reduce graft rejection episodes, it has been proposed that adherence to IST should be over 97.0%, and only one dose in a month could be missed out. (27) Missing doses could affect tacrolimus C_trough_ level. Our study found no significant association between adherence level and marital status, occupation, education level, and length of time since kidney transplantation. However, a direct link between the length of time since transplantation and nonadherence was reported in some studies.(13) Furthermore, older patients had a higher adherence rate than younger patients (29) with more nonadherence observed in a single individual. (15) Some studies reported higher adherence rates in professionals compared with unskilled employees, while other studies reported no difference in socioeconomic class or educational attainment. (13)As suggested, sociodemography factors affecting adherence rate are subjective (30) and may not be the factor of nonadherence in Bhutan as the states take care of health expenses, including long-term maintenance of IST. Only 8 (25.8%) individuals took a lower dose of IST than the prescribed dose. Monitoring of tacrolimus C_trough_ levels is, on the other hand, the objective index to determine drug exposure currently used in routine monitoring as part of patient care after transplantation. Seventy-two percent were found to have tacrolimus C_trough_ level variation above 20% and 39 (67.2%) identified as nonadherence on clinician assessment. There was no significant difference (P<0.001) between the clinician’s assessment score and tacrolimus level. Overall, the %CV of ≤ 20 % in 16 of 58 patients (27.5%) was consistent with good adherence. Those with %CV between 20-43% in 33 of 58 patients (57.0%) and %CV ≥ 43% in 9 of 58 patients (15.5%) were identified as medium and poor adherence, respectively. The medium and poor adherence KTRs had raised serum creatinine and BUN, which was in agreement with a previous study in Germany and Austria. (25) The current study showed that clinicians’ assessment of adherence rate was correlated with the %CV of tacrolimus C_trough_ level, indicating that tacrolimus C_trough_ level is applicable in identifying nonadherence in routine clinical care.

## Conclusion

The assessment of adherence to immunosuppressive therapy provides information on factors linked to nonadherence. Kidney transplant recipients concerned about graft survival are more adhering. The nonadherent patient requires close observation and frequent monitoring of immunosuppressive levels.

## List of abbreviations

IST: Immunosuppressive therapy
KTRs: Kidney transplant recipients
QoL: Quality of life
BAASIS: Basel Assessment of Adherence to immunosuppressive medication scale
TDM: Therapeutic drug monitoring
AUC: Area Under the Concentration
C _trough_: Concentration trough level
CV%: Percentage covariate of variation

## Declarations

## Acknowledgement

We would like to acknowledge the management and clinical staff of Jigme Dorji Wangchuk National Referral Hospital for their support during recruitment and data collection. The staff of the biochemistry unit and TDM unit helped in performing drug-level monitoring. We would like to acknowledge them for their continuous support.

## Ethical approval

This study was performed in line with the principles of the Declaration of Helsinki. Approval was granted by the Ethics Committee of the Research Ethics Board of Health of Bhutan (ref. REBH/Approval/2021/077), the Ministry of Health (ref. PPD/admin. Cl/(9)/2020-21/100) and the management of Jigme Dorji Wangchuk national referral hospital with reference to PPD/admin. Cl/(9)/2020-21/100 and JDWNRH/MERU/01/2020 - 2021/10054.

## Consent to participate and publication

Written informed consent was obtained from individuals to participate and for the publication of this study. Informed consent translated both into English and Dzongkha (Bhutan national language) was available to the participants.

## Authors’ contributions

All authors contributed to the study’s conception and design. Material preparation, data collection, and analysis were performed by Choki Dorji and Kesara Na-Bangchang. The first draft of the manuscript was written by Choki Dorji, and all authors commented on previous versions of the manuscript. All authors read and approved the final manuscript.

## Author’s information and affiliations

Mr Choki Dorji is a clinical pharmacist at Jigme Dorji Wangchuk national referral hospital in Bhutan and has a master’s degree in clinical Pharmacy. He has a working experience as a transplant pharmacist and therapeutic drug monitoring of immunosuppressants.

## Data Availability

- Tables 1, 2, 3 and 4 attached
- Figure 1 is attached.
- The translated version of the data collection tool is attached as a supplementary file.
- The datasets used and/or analysed during the current study are available from the corresponding author upon reasonable request.

## REFERENCES

1. Shao Y, Fan Y, Xie Y, Yin L, Zhang Y, Deng L, et al. Effect of continuous renal replacement therapy on kidney injury molecule-1 and neutrophil gelatinase-associated lipocalin in patients with septic acute kidney injury. Exp Ther Med. 2017 Jun 1;13(6):3594.

2. Jay S, Litt IF, Durant RH. Compliance with therapeutic regimens. Journal of Adolescent Health Care. 1984 Apr 1;5(2):124–36.

3. Dew MA, DiMartini AF, de Vito Dabbs A, Myaskovsky L, Steel J, Unruh M, et al. Rates and risk factors for nonadherence to the medical regimen after adult solid organ transplantation. Transplantation. 2007;83(7):858–73.

4. Eduardo Sabaté. Adherence to long-term therapies: evidence for action, World Health Organization, Geneva, Switzerland, 2003. [Internet]. [cited 2023 Feb 13]. Available from: https://apps.who.int/iris/handle/10665/42682

5. Chisholm-Burns M, Pinsky B, Parker G, Johnson P, Arcona S, Buzinec P, et al. Factors related to immunosuppressant medication adherence in renal transplant recipients. Clin Transplant. 2012;26(5):706–13.

6. Pinsky BW, Takemoto SK, Lentine KL, Burroughs TE, Schnitzler MA, Salvalaggio PR. Transplant outcomes and economic costs associated with patient noncompliance to immunosuppression. American Journal of Transplantation. 2009;9(11):2597–606.

7. Jindel RM, Joseph JT, Morris MC, Santella RN, Baines LS. Noncompliance after kidney transplantation: A systematic review. Transplant Proc. 2003;35(8):2868–72.

8. Butler JA, Peveler RC, Roderick P, Horne R, Mason JC. Measuring compliance with drug regimens after renal transplantation: Comparison of self-report and clinician rating with electronic monitoring. Vol. 77, Transplantation. Transplantation; 2004. p. 786–9.

9. Sellarés J, De Freitas DG, Mengel M, Reeve J, Einecke G, Sis B, et al. Understanding the causes of kidney transplant failure: The dominant role of antibody-mediated rejection and nonadherence. American Journal of Transplantation. 2012 Feb;12(2):388–99.

10. Fine RN, Becker Y, De Geest S, Eisen H, Ettenger R, Evans R, et al. Nonadherence consensus conference summary report. Am J Transplant. 2009 Jan;9(1):35–41.

11. WHO. Adherence to Long-Term Therapies: Evidence for Action, 2003 - PAHO/WHO | Pan American Health Organization [Internet]. [cited 2023 May 19]. Available from: https://www.paho.org/en/documents/who-adherence-long-term-therapies-evidence-action-2003

12. Gaynor JJ, Guerra G, Roth D, Chen L, Kupin W, Mattiazzi A, et al. Graft failure due to nonadherence among 150 prospectively-followed kidney transplant recipients at 18 years post-transplant: Our results and review of the literature. J Clin Med. 2022 Feb 28;11(5).

13. Greenstein S, Siegal B. Odds probabilities of compliance and noncompliance in patients with a functioning renal transplant: a multicenter study. Transplant Proc. 1999 Feb;31(1– 2):280–1.

14. Kiley DJ, Lam CS, Pollak R. A study of treatment compliance following kidney transplantation. Transplantation. 1993;55(1):51–6.

15. De Geest S, Borgermans L, Gemoets H, Abraham I, Vlaminck H, Evers G, et al. Incidence, determinants, and consequences of subclinical noncompliance with immunosuppressive therapy in renal transplant recipients. Transplantation. 1995;59(3):340–7.

16. Kalil RSN, Heim-Duthoy KL, Kasiske BL. Patients with a low income have reduced renal allograft survival. Am J Kidney Dis. 1992;20(1):63–9.

17. De Geest S, Dobbels F, Fluri C, Paris W, Troosters T. Adherence to the therapeutic regimen in heart, lung, and heart-lung transplant recipients. J Cardiovasc Nurs. 2005;20(5 Suppl).

18. Hilbrands LB, Hoitsma AJ, Koene RAP. Medication compliance after renal transplantation. Transplantation. 1995 Nov 15;60(9):914–20.

19. Cukor D, Ver Halen N, Pencille M, Tedla F, Salifu M. A Pilot Randomized Controlled Trial to Promote Immunosuppressant Adherence in Adult Kidney Transplant Recipients. Nephron. 2017;135(1):6–14.

20. Belaiche S, Décaudin B, Dharancy S, Noel C, Odou P, Hazzan M. Factors relevant to medication non-adherence in kidney transplant: a systematic review. Int J Clin Pharm. 2017 Jun 1;39(3):582–93.

21. Cramer JA, Mattson RH, Prevey ML, Scheyer RD, Ouellette VL. How often is medication taken as prescribed? A novel assessment technique. JAMA. 1989 Jun 9;261(22):3273–7.

22. Dorji C, Tobgay T, Na-Bangchang K. Title: The factors affecting the HRQOL of kidney transplant recipients in the land of Gross National Happiness, Bhutan - A cross-sectional study. 2023 May 11 [cited 2023 Jul 31]; Available from: 10.21203/rs.3.rs-2857908/v1

23. Marsicano EDO, Fernandes NDS, Colugnati F, Grincenkov FRDS, Fernandes NMDS, De Geest S, et al. Transcultural adaptation and initial validation of Brazilian-Portuguese version of the Basel assessment of adherence to immunosuppressive medications scale (BAASIS) in kidney transplants. BMC Nephrol. 2013;14(1).

24. Akchurin OM, Melamed ML, Hashim BL, Kaskel FJ, Rio M Del. Medication Adherence in the Transition of Adolescent Kidney Transplant Recipients to the Adult Care. Pediatr Transplant. 2014;18(5):538–48.

25. Kreuzer M, Prüfe J, Oldhafer M, Bethe D, Dierks ML, Müther S, et al. Transitional care and adherence of adolescents and young adults after kidney transplantation in Germany and Austria. Medicine (United States). 2015;94(48):1–8.

26. Thinley S, Tshering P, Wangmo K, Wangmo K, Wangchuk N, et al. Health systems in transition. 2017. The kingdom of Bhutan health system review.

27. Ganjali R, Sabbagh MG, Nazemiyan F, Mamdouhi F, Aval SB, Taherzadeh Z, et al. Factors Associated With Adherence To Immunosuppressive Therapy And Barriers In Asian Kidney Transplant Recipients. Immunotargets Ther. 2019 Nov 7;8:53–62.

28. Moreso F, Torres IB, Costa-Requena G, Serón D. Nonadherence to immunosuppression: challenges and solutions. Transplant Research and Risk Management. 2015 Jun 3;7:27–34.

29. Schweizer RT, Rovelli M, Palmeri D, Vossler E, Hull D, Bartus S. Noncompliance in organ transplant recipients. Transplantation. 1990;49(2):374–7.

30. Raiz LR, Kilty KM, Henry ML, Ferguson RM. Medication compliance following renal transplantation. Transplantation. 1999 Jul 15;68(1):51–5.

